# A novel diagnostic intracranial EEG biomarker in MOGHE

**DOI:** 10.64898/2026.06.05.26355018

**Authors:** Vadym Gnatkovsky, Ekaterina Poguzhelskaya, Valeri Borger, Rainer Surges, Kerstin Alexandra Klotz, Valentina Zschernack, Till Hartlieb, Manfred Kudernatsch, Ahmed Gaballa, Thomas Cloppenborg, Friedrich G. Woermann, Thilo Kalbhenn, Hajo Hamer, Stephanie Gollwitzer, Stefan Rampp, Daniel Delev, Florian Mayer, Karl Rössler, Valerie Anne Quinot, Angelika Mühlebner, Rafael Toledano, Antonio Gil-Nagel, Roland Coras, Ingmar Blümcke, Katja Kobow

**Affiliations:** Department of Epileptology, University Hospital Bonn, Bonn, Germany, Partner of the ERN EpiCARE; Department of Neurosurgery, University Hospital Bonn, Bonn, Germany, Partner of the ERN EpiCARE; Department of Neuropediatrics, University Hospital Bonn, Bonn, Germany, Partner of the ERN EpiCARE; Institute for Neuropathology, University Hospital Bonn, Bonn, Germany, Partner of the ERN EpiCARE; Center for Pediatric Neurology, Neurorehabilitation, and Epileptology, Schön Clinic Vogtareuth, Vogtareuth, Germany; Center for Neurosurgery and Epilepsy Surgery, Schön Clinic Vogtareuth, Vogtareuth, Germany; Research Institute „Rehabilitation, Transition, Palliation”, PMU Salzburg, Salzburg, Austria; Department of Epileptology, Krankenhaus Mara, Bethel Epilepsy Centre, Bielefeld University, Campus Bielefeld-Bethel, Bielefeld, Germany; Department of Neurosurgery, Medical School OWL, Bielefeld University, Campus Bielefeld-Bethel, Bielefeld, Germany; Department of Neurology, Epilepsy Center, Universitätsklinikum Erlangen, Friedrich-Alexander University Erlangen-Nürnberg (FAU), Erlangen, Germany, Partner of the ERN EpiCARE; Department of Neuroradiology, Universitätsklinikum Erlangen, Friedrich-Alexander University Erlangen-Nürnberg (FAU), Erlangen, Germany, Partner of the ERN EpiCARE; Department of Neurosurgery, Universitätsklinikum Erlangen, Friedrich-Alexander University Erlangen-Nürnberg (FAU), Erlangen, Germany, Partner of the ERN EpiCARE; Department of Neuropathology, Universitätsklinikum Erlangen, Friedrich-Alexander University Erlangen-Nürnberg (FAU), Erlangen, Germany, Partner of the ERN EpiCARE; Department of Pediatrics and Adolescent Medicine, Medical University Vienna, Vienna, Austria. Partner of the ERN EpiCARE; 6bDepartment of Neurosurgery, Medical University Vienna, Vienna, Austria. Partner of the ERN EpiCARE; Division of Neuropathology and Neurochemistry, Department of Neurology, Medical University Vienna, Vienna, Austria. Partner of the ERN EpiCARE; Department of Pathology, University Medical Center Utrecht, Utrecht, The Netherlands, Partner of the ERN EpiCARE; Department of Neurology, Epilepsy Center, Hospital Ruber Internacional, Madrid, Spain, Partner of the ERN EpiCARE

## Abstract

Mild malformation of cortical development with oligodendroglial hyperplasia and epilepsy (MOGHE) is a recently recognized cause of drug-resistant focal epilepsy. It is often MRI-negative or shows imaging features mimicking focal cortical dysplasias, which makes recognition difficult and limits presurgical counseling. We aimed to identify an intracranial EEG (iEEG) biomarker that distinguishes MOGHE from other developmental brain lesions encountered in epilepsy surgery. In a retrospective multicenter test cohort of 38 patients (18 MOGHE, 20 non-MOGHE), we analyzed long-term stereo-EEG and subdural recordings. Only MOGHE patients showed highly stereotyped clusters of very brief low-voltage fast activity (LVFA) events, organized into status-like 3 to 12-minute episodes that often lacked clear clinical symptoms. LVFA clusters were present in 16/18 MOGHE and 0/22 non-MOGHE patients. We then tested diagnostic performance in an independent, blinded single-center validation cohort of 22 patients (11 MOGHE, 11 non-MOGHE), in which visual identification of LVFA clusters correctly classified 10/11 MOGHE and 10/11 non-MOGHE cases (Cohen’s κ=0.82). Penalized logistic regression further confirmed MOGHE histology as the strongest predictor of LVFA clusters, independent of age and lobe localization. Because LVFA clusters can be recognized visually on routine intracranial EEG recordings without specialized software, this biomarker is readily applicable in clinical practice and may improve presurgical identification of MOGHE. Future prospective studies should determine whether its recognition influences surgical planning, improves outcome prediction, or facilitates selection of patients for mechanism-based therapies.

**Highlights:**

- Clusters of brief LVFA in intracranial EEG are a lesion-specific biomarker of MOGHE compared to mMCD and FCD2
- LVFA clusters reliably distinguished MOGHE from non-MOGHE lesions (mMCD and FCD2) in both a multicenter test cohort and an independent blinded validation cohort.
- Younger age at seizure onset is a secondary independent modifier, while frontal localization and age at intracranial recording are not.
- Clinicians can detect LVFA clusters on continuous intracranial EEG using routine montages and ≥60-second review windows, without any specialized software.
- LVFA clusters show a characteristic temporal organization and burden in MOGHE, indicating a distinct pattern of pathological network activity in this lesion type.

## Introduction

MOGHE is a rare, recently recognized developmental brain malformation. It is often associated with severe pediatric focal-onset epilepsy and other neurodevelopmental comorbidities, affecting both males and females ^1–3^. Histopathologically, MOGHE is characterized by the misplacement of ectopic neurons in the white matter and increased oligodendroglial cell densities alongside a yet-to-be-specified myelination defect ^4; 5^. Two main clinical phenotypes have been described in MOGHE, including early epileptic encephalopathy with epileptic spasms as the predominant seizure type and intellectual disability, and a milder drug-resistant focal epilepsy associated with relatively preserved cognition ^1^.

MRI findings in MOGHE often resemble focal cortical dysplasia and include cortical thickening, blurring of the grey-white matter junction ^1; 2; 5–7^. Laminar signal hyperintensities at the grey-white matter junction or reduced signal contrast between grey and white matter have also been described, appear to be age-related and subject to change with brain maturation, particularly myelination ^4; 8^. MOGHE predominantly localizes in frontal lobes but can be challenging to delineate on MRI, sometimes presenting as an extended lesion covering multiple lobes. Post-surgical outcomes regarding seizure freedom may vary significantly, depending on surgical strategy (e.g., subtotal or partial lobectomy versus lesionectomy), disease duration, and molecular-genetic factors ^1; 2; 5–8^. However, cognitive performance remains unchanged in most patients after surgery ^1^.

Up to 50% of patients with MOGHE harbor brain-somatic variants in the galactose transporter gene SLC35A2 ^1; 9–11^, and recent studies have additionally identified frequent somatic Y-chromosome gain within lesional white matter, highlighting the molecular heterogeneity of the disease ^12^. Germline SLC35A2 variants cause a congenital disorder of glycosylation associated with developmental and epileptic encephalopathy ^13^, and oral galactose supplementation has shown beneficial effects in both SLC35A2-related disease and selected MOGHE patients ^9; 14^. In addition, disease-specific DNA methylation signatures have emerged as promising diagnostic biomarkers that may complement conventional histopathological classification ^15–17^.

Overall, MOGHE remains underrecognized during presurgical evaluation. MRI abnormalities are often subtle, age-dependent, and difficult to delineate, whereas definitive diagnosis typically relies on expert histopathological examination after surgery ^16^. Surgical outcome appears to depend on lesion extent and resection strategy, with incomplete resections associated with less favorable seizure outcomes in some cohorts ^3^. Consequently, accurate preoperative identification of MOGHE may have direct implications for the interpretation of intracranial EEG findings, delineation of the epileptogenic zone, and surgical planning.

Intracranial EEG, particularly stereo-EEG, is frequently used in patients with suspected MOGHE because lesion boundaries are often ill-defined and may extend across multiple lobes and beyond the apparent MRI abnormality. However, no intracranial EEG biomarker has yet been established for reliable recognition of MOGHE during presurgical evaluation. Such a biomarker could improve etiological diagnosis before surgery, support interpretation of widespread or atypical epileptic networks, guide resection strategies, and facilitate patient selection for emerging molecularly informed therapies, including SLC35A2-targeted approaches.

Here, we hypothesized that intracranial EEG contains disease-specific features that may facilitate presurgical recognition and stratification of MOGHE. Using qualitative and quantitative analyses of intracranial EEG recordings, we identified a characteristic electrophysiological pattern associated with MOGHE. To assess diagnostic specificity and clinical utility, we performed an independent evaluator-blinded validation study that included MOGHE and non-MOGHE cases. Our findings establish a novel intracranial EEG biomarker that may improve presurgical recognition and stratification of MOGHE and provide additional information relevant to surgical decision-making.

## Methods

### Cohort description

This study, including the retrospective audit of EEG data collected during standard clinical care, was approved by the Ethics Committees of the Medical Faculties of Friedrich-Alexander-Universität Erlangen-Nürnberg, Germany (AZ 92_14B, 193_18B, and 18-193_1-Bio), Rheinische Friedrich-Wilhelms-Universität Bonn (No. 352/12, 404/17), Medical University Vienna (1357/2021). Written informed consent for the research use of clinical and molecular genetic data was obtained from all patients or, in cases of intellectual disability, from their parents or legal guardians.

The aim of this multicenter retrospective study was to identify an intracranial EEG biomarker that distinguishes patients with focal epilepsy due to mild malformation of cortical development with oligodendroglial hyperplasia in epilepsy (MOGHE) from patients with focal epilepsy of non-MOGHE etiology. Because MOGHE is a rare lesion and intracranial EEG is available only in selected presurgical cases, the sample size was determined by the availability of histopathologically confirmed cases with analyzable intracranial EEG recordings and centralized histopathological review (IB, RB) according to current international classification criteria ^18^. The primary diagnostic endpoint was the presence or absence of the diagnostic biomarker; accordingly. Diagnostic performance was evaluated using sensitivity, specificity, positive and negative predictive values, Fisher’s exact tests, Cohen’s κ, and penalized logistic regression.

The study comprised a test cohort (n=38) and an independent validation cohort (n=22). The test cohort included 18 histologically defined MOGHE patients (8 female, 10 male) and 20 non-MOGHE patients diagnosed with either mild malformation of cortical development (mMCD; n=9) or focal cortical dysplasia type II (FCD2; n=11). In MOGHE, the age at epilepsy onset was marked earlier (median 3.5 years, IQR 1.25-6.75) compared with non-MOGHE (median 7.5 years, IQR 5.0-13.25; Mann-Whitney U test, *p*=0.003), as was age at intracranial EEG implantation in MOGHE (median 12.0 years, IQR 6.25-20.75) compared to non-MOGHE (24.5 years, IQR 16.75-27.25; *p*=0.007).

The independent validation cohort comprised 22 patients (9 female, 13 male) with histopathologically confirmed MOGHE (n=11), mMCD (n=4), or FCD2 (n=7). In this cohort, age at epilepsy onset was again lower in MOGHE (median 2.0 years, IQR 0.81-3.05) than in non-MOGHE (6.58 years, IQR 5.25-11.50; *p*=0.0006), as was age at intracranial EEG implantation (MOGHE: 6.44 years, IQR 5.28-12.48; non-MOGHE: 21.54 years, IQR 16.69-34.39; *p*=0.006). There was no clinical difference between cohorts (**Table 1**).

**Table 1:** Summary of test and validation cohorts.

| Variable | Test (n=38) | Validation (n=22) | Test | p-value |
| --- | --- | --- | --- | --- |
| Age at onset | 6.0 [3.1–10.8] | 4.0 [2.0–7.2] | Wilcoxon rank-sum | 0.173 |
| Age at iEEG | 19.0 [11.0–25.8] | 15.4 [6.9–29.2] | Wilcoxon rank-sum | 0.753 |
| MOGHE/non-MOGHE | 18 (47%) / 20 (53%) | 11(50%) / 11 (50%) | Fisher's exact | 1.000 |
| Sex (f/m) | 18 (47%) / 20 (53%) | 9 (41%) / 13 (59%) | Fisher's exact | 0.789 |
| LVFA cluster (Y/N) | 16 (42%) / 22 (58%) | 11 (50%) / 11 (50%) | Fisher's exact | 0.599 |
| Localization (frontal/other) | 23 (61%) /15 (39%) | 16 (73%) / 6 (27%) | Fisher's exact | 0.408 |

All patients had drug-resistant focal epilepsy and underwent phase II presurgical evaluation with intracranial EEG. Depending on local clinical practice and indication, recordings were obtained using either subdural grid and strip electrodes or 5-18 multichannel depth electrodes (Ad-Tech Medical, Oak Creek, USA, or DIXI Medical, Besançon, France). Depth electrodes had diameters of 0.8-1.2 mm and contained 5-18 contacts with 1.5-2 mm intercontact spacing. Electrode implantation was planned to sample the presumed epileptogenic network based on clinical findings, seizure semiology, ictal EEG, and neuroimaging results. Continuous intracranial EEG recordings were acquired over 4-15 days using different recording systems, including Nihon Kohden (Rosbach, Germany), Sienna (EMS Biomedical, Austria), Micromed/Natus (Middleton, USA), and Compumedics (Freiberg, Germany). Recordings were obtained with a band-pass filter of 0.016-300 Hz and a sampling rate of 0.256-1 kHz. Multiple seizures and interictal events were recorded in all patients. For analysis, either representative intracranial EEG segments over several hours or, whenever available, the full continuous recordings were reviewed.

MRI-visible lesions were reported in 83% of cases (50/60), most commonly FCD-like abnormalities, including cortical thickening, blurring of the gray-white matter junction, and FLAIR hyperintensity. In patients with MOGHE, lesions were predominantly located in the frontal lobe, whereas in the non-MOGHE group, lesions were more heterogeneously distributed across frontal, temporal, and parietal regions (**Table 1**). FDG-PET, was not systematically performed, but showed focal hypometabolism in 25/30 patients concordant with the presumed epileptogenic region, particularly in cases with subtle or nonspecific MRI abnormalities.

Genetic testing was performed as part of routine clinical care in 29 of 60 patients (48%) using array comparative genomic hybridization (array-CGH), fluorescence in situ hybridization (FISH), targeted gene panels, or whole-exome sequencing. No systematic molecular analysis of resected brain tissue and matched blood samples was performed. A molecular diagnosis was provided in 10 out of 29 tested patients (34%), including six MOGHE patients with brain-somatic *SLC35A2* variants or Y-chromosome gain, one mMCD patient with a *SLC6A5* variant, and three FCD2A patients with germline variants in *NPRL3* and *DEPDC5*.

More detailed clinical descriptions of 28/29 MOGHE patients included in this study, including seizure semiology, genetics, and imaging findings, have been provided previously ^1; 2^.

### Qualitative intracranial EEG analysis

Heterogeneous multicenter stereo-EEG datasets were converted and reviewed using Persyst EEG software (Solana Beach, USA). Subsequent quantitative analyses were performed using the previously described LabVIEW-based Elpho platform (www.elpho.it; National Instruments, USA) ^19^. This tool supports real-time extraction and visualization of intracranial EEG biomarkers within the epileptic network, focusing on spectral dynamics and slow wave components of individual contacts. Computer-assisted analysis was applied to all recorded leads. Electrographic biomarkers and time-frequency profiles were quantified in full-length recordings when available or in representative ictal and interictal recording segments per patient, following established procedures ^19–21^. Briefly, fast activity power (80-130Hz) was calculated for each contact and seizure. Contacts with values exceeding the 95%-confidence interval across all channels were visually reviewed to identify low-voltage fast activity (LVFA) and “chirps” (oscillatory patterns with decreasing frequency). Events followed by tonic spiking, bursting, and postictal depression were classified as electrographic seizures, irrespective of the presence or absence of clinical manifestations (e.g., tonic seizures, epileptic spasms). Conversely, clinical events without a corresponding electrographic seizure pattern were not counted as seizures. LVFA events outside of typical electrographic seizures were also analyzed. Extended time windows (1-5 min) were used to assess temporal dynamics and identify clustering.

In total, 150.2 days of intracranial EEG were reviewed (mean 3.8 days per patient; range 0.15-10.5 days), and mean seizure frequency (seizures/week) per patient was calculated for group comparison (**Table 1**). Data of test and validation cohorts were evaluated independently by a single reviewer (VG), who was blinded to clinical information, histopathological diagnoses, the number of MOGHE cases, and all imaging data and genetic findings in the validation cohort.

### Quantitative intracranial EEG analysis

In 28 patients (18 MOGHE, 10 non-MOGHE), we performed quantitative analysis of continuous intracranial EEG recordings. Data, originally stored in various proprietary formats, were converted to binary files using Persyst software (Persyst Development LLC, V14), and all subsequent analyses were performed in MATLAB (The MathWorks, R2024b, version 24.2.0.2806996). Channels with marked high-frequency artifacts (e.g., muscle activity) were excluded, and all remaining data were analyzed using a bipolar montage. Given the long recording durations, data were segmented into batches of 0.5-3 hours, depending on sampling rate and channel count.

Raw signals were screened for high-amplitude artifacts using a moving median-based outlier detection method (120 s sliding window, threshold 15x scaled median absolute deviation). Artifact segments were attenuated using a smooth 5 s Hann-window transition mask to preserve temporal continuity. Each batch was then filtered between 60 and 120 Hz (zero-phase band-pass), and the root-mean-square (RMS) envelope of the filtered signal was computed with a 200 ms sliding window. Channels with excessive variance were excluded using an outlier criterion (12x standard deviation across channels). For the remaining channels, a common inter-channel projection over time was estimated and subtracted to suppress baseline activity and shared artifacts.

For LVFA event detection, channel-specific amplitude thresholds were derived from the scaled median absolute deviation of each batch. Peaks in the RMS envelope exceeding these thresholds were identified, and events were defined by onset and offset at 15% of peak amplitude above baseline. Overlapping or closely spaced peaks (<150 ms apart) were merged, and nested events were collapsed to a single event with the highest peak. To suppress ringing artifacts from sharp transients, we quantified the number of peaks in the unfiltered signal during each candidate event and discarded events with a peak rate <50 Hz. Detected events and recording segments were visually reviewed on a per-patient basis, and channels dominated by persistent noise were excluded. For each validated LVFA event, we extracted peak amplitude, duration, onset time, and channel location.

To identify LVFA clusters, we applied a custom inter-event-interval-based algorithm. Within each channel, a cluster was defined as a sequence of at least 10 LVFA events with inter-event intervals generally <15 s and a minimum separation of 2 s between events. Seizure counts and durations were obtained from clinical records or, when unavailable, by expert review and annotation of the recordings.

To explore temporal organization, we examined the occurrence of LVFA events and clusters across the 24-h cycle. For each patient, LVFA event and cluster onsets were mapped to clock time, pooled, and visualized as hourly histograms to assess diurnal patterns and potential circadian modulation of LVFA clustering. We further summarized recurrent cluster onsets in a circular (24-h) plot and computed the mean resultant vector, displaying its peak angle at the corresponding time of day as an estimate of preferred LVFA event and cluster phase.

### Statistics

Statistical analyses were performed using GraphPad Prism (GraphPad Software, Boston, MA, USA) and Python. Continuous variables were compared between groups using two-sided Mann-Whitney U (Wilcoxon rank-sum) tests, consistent with non-normal distributions and small group sizes, and categorical frequencies, including clustered LVFA prevalence, using Fisher’s exact test. Clustered LVFA was modeled as a binary outcome in the combined cohort, with histology dichotomized as MOGHE versus non-MOGHE and localization as frontal versus non-frontal. Given sparse cells and quasi-complete separation, the primary multivariable model was a ridge-penalized logistic regression adjusting for histology, localization, age at seizure onset, and age at iEEG implantation; continuous predictors were standardized, and the penalty was tuned by 10-fold cross-validation. Coefficient uncertainty was quantified by 1,000 bootstrap resamples, and discrimination by apparent and cross-validated ROC AUC. Firth bias-reduced logistic regression with the same covariates served as a sensitivity analysis. Prespecified frontal-only and pediatric-only subgroup analyses used Fisher’s exact test with continuity-corrected odds ratios and 95% confidence intervals. All tests were two-sided; p < 0.05 was considered significant.

## Results

### Test cohort: A novel qualitative iEEG pattern with repetitive clusters of LVFA characterizes MOGHE

This study aimed to identify intracranial EEG (iEEG) features that could support the diagnosis and clinical management of MOGHE. We first analyzed a retrospective test cohort of 38 patients: 18 with histopathologically confirmed MOGHE and 20 non-MOGHE cases with mild malformations of cortical development (mMCD), focal cortical dysplasia type 2 (FCD2), as well as one case with a complex malformation including FCD2A and polymicrogyria. We selected these comparison groups because, like MOGHE, they are often difficult to recognize in routine clinical practice before surgery, due to absent or non-specific MRI findings and the lack of established iEEG markers.

We examined raw, unfiltered iEEG recordings and corresponding spectral features from a total of 150.2 recording days, including ictal and interictal segments in both MOGHE (**Fig. 1**) and non-MOGHE (**Fig. 2**). Across the 38 patients, we analyzed 184 focal electrographic seizures. Seizure duration typically ranged from 20 to 60 seconds. Seizure onset was consistently marked by low-voltage fast activity (LVFA) in the 80-130 Hz range, appearing as a distinct “chirp-like” pattern in the power spectrogram and followed by tonic spiking, a bursting phase, and postictal depression (**Fig. 1A, 2A**). Most seizure onsets arose from the frontal lobe (n = 19), with fewer cases involving the temporal (n = 6) or other lobes (n = 3; **Table 1**). Median seizure burden did not differ significantly between MOGHE and non-MOGHE patients (MOGHE = 11.8 seizures/week, range 0-61; non-MOGHE = 14.0 seizures/week, range 0-47; Mann-Whitney U-test, *p*>0.05).

**Figure 1:**
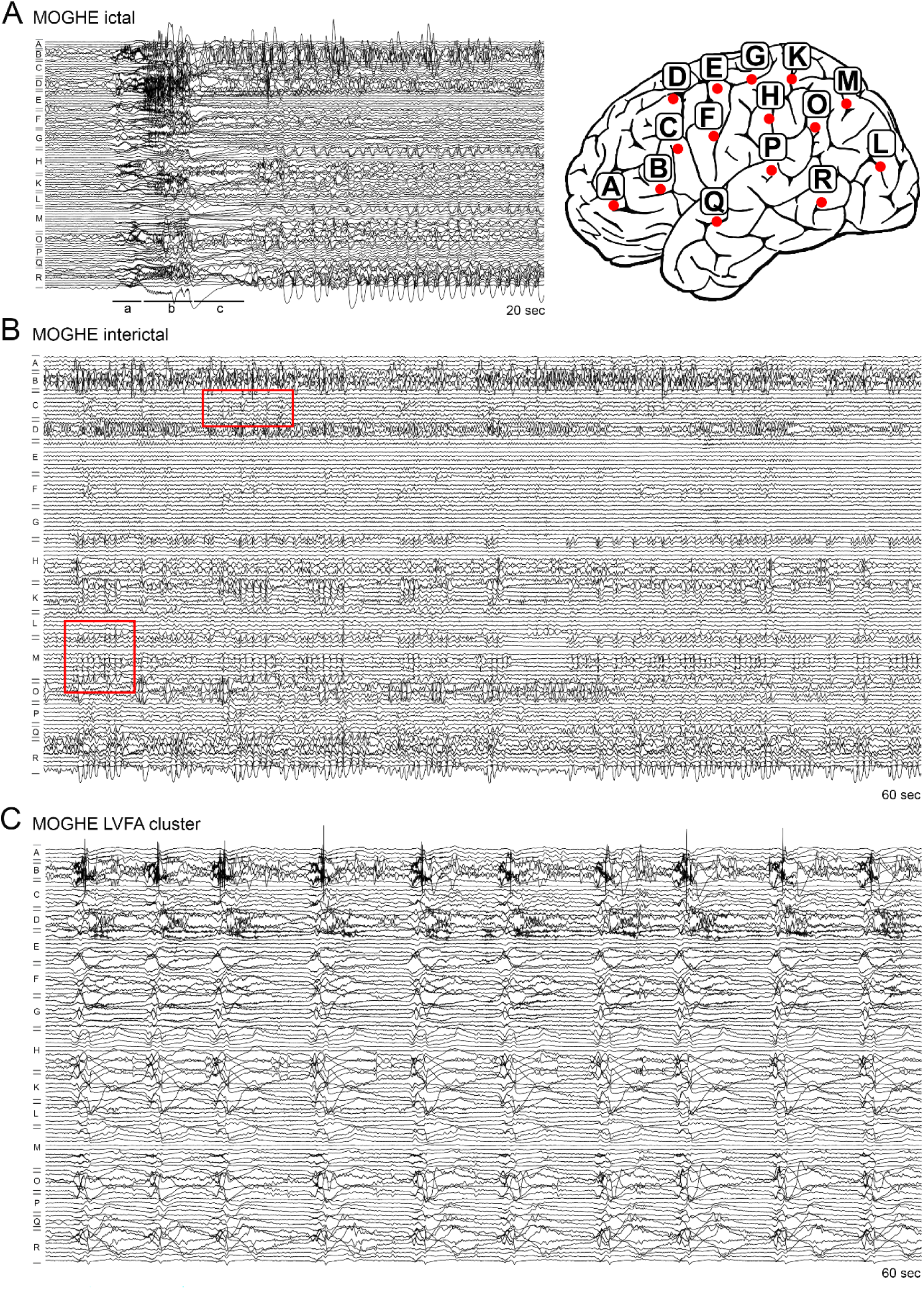
MOGHE iEEG characteristics. (**A**) Representative ictal iEEG pattern in MOGHE showing focal seizure onset characterized by LVFA (a), followed by progression into tonic spiking discharges and a bursting phase (b), and postictal depression (c; left panel). Corresponding implantation scheme of the same patient demonstrating depth electrode coverage of the frontal, temporal, and parietal lobes (right panel). (**B**) Representative interictal iEEG activity demonstrating diffuse rhythmic spiking across multiple electrode contacts (red rectangles). (**C**) Sixty-second intracranial EEG segment illustrating a characteristic LVFA cluster with recurrent rhythmic LVFA events occurring at a frequency of approximately 10 events/min.

**Figure 2:**
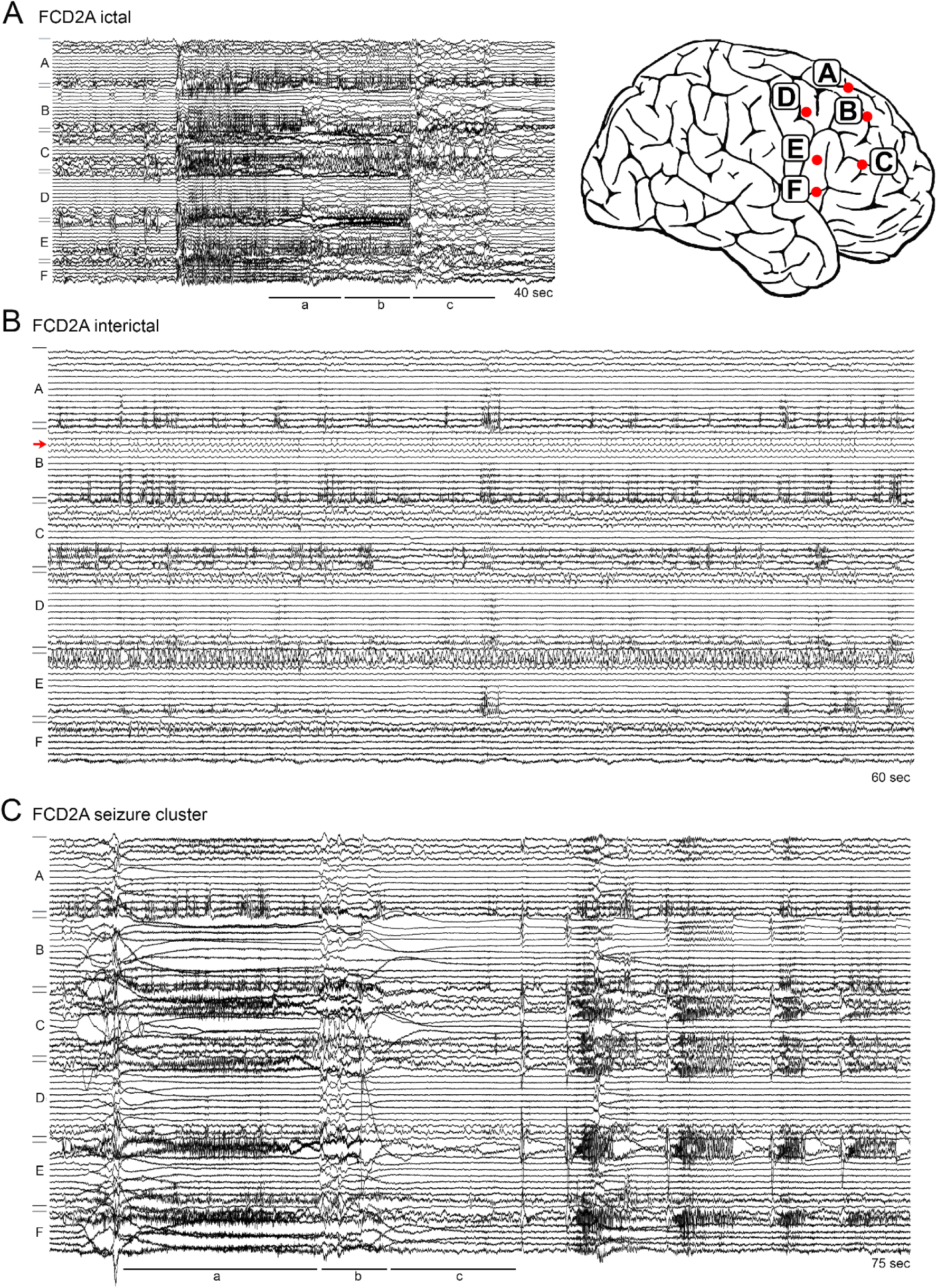
Non-MOGHE iEEG characteristics. (**A**) Representative ictal iEEG pattern in FCD2A showing a focal seizure onset characterized by LVFA (a), followed by progression into tonic spiking discharges and a bursting phase (b), and postictal depression (c; left panel). Corresponding implantation scheme of the same patient demonstrating depth electrode coverage of the frontal lobe (right panel); the contralateral implantation is omitted for clarity. (**B**) Representative interictal iEEG activity demonstrating focal rhythmic spiking involving a limited number of contacts (red arrow). (**C**) Intracranial EEG segment illustrating a focal seizure cluster in FCD2A comprising five seizures within 75 seconds. The initial seizure is followed by several additional brief seizures (<10 s). Despite their short duration, all events exhibit the characteristic electrographic seizure sequence, including LVFA at onset (a), bursting activity (b), and postictal depression (c).

Outside of electrographic seizures, both MOGHE and non-MOGHE patients showed continuous rhythmic spiking at 1-2 Hz in the same anatomical regions as seizure onset (**Fig. 1B, 2B**). In MOGHE, however, interictal spiking was more diffuse and involved a larger number of contacts and electrodes than is typically observed in FCD2.

Only MOGHE patients in the test cohort exhibited a striking and previously unrecognized iEEG pattern consisting of repetitive, very brief LVFA events lasting 1-2 seconds and recurring at a rate of 4-11 events per minute (**Figs. 1C, 2C**). These events did not evolve into full seizures and were not followed by synchronized bursting and postictal depression. Instead, they occurred in highly rhythmic clusters lasting 3-12 minutes with high intra- and interindividual variability, which were most readily identified when recordings were reviewed over longer time windows (>60 s) and were observed on a daily basis (**Fig. 3A-B**). During these clusters, a substantial proportion of the sampled epileptic network was progressively recruited, and the frequency of LVFA events gradually decreased over the course of each cluster, indicating reproducible internal dynamics (**Fig. 3B-C**). Electrographically, the prolonged duration, spatial extent, and evolving structure of these clusters gave them a status-like appearance.

**Figure 3:**
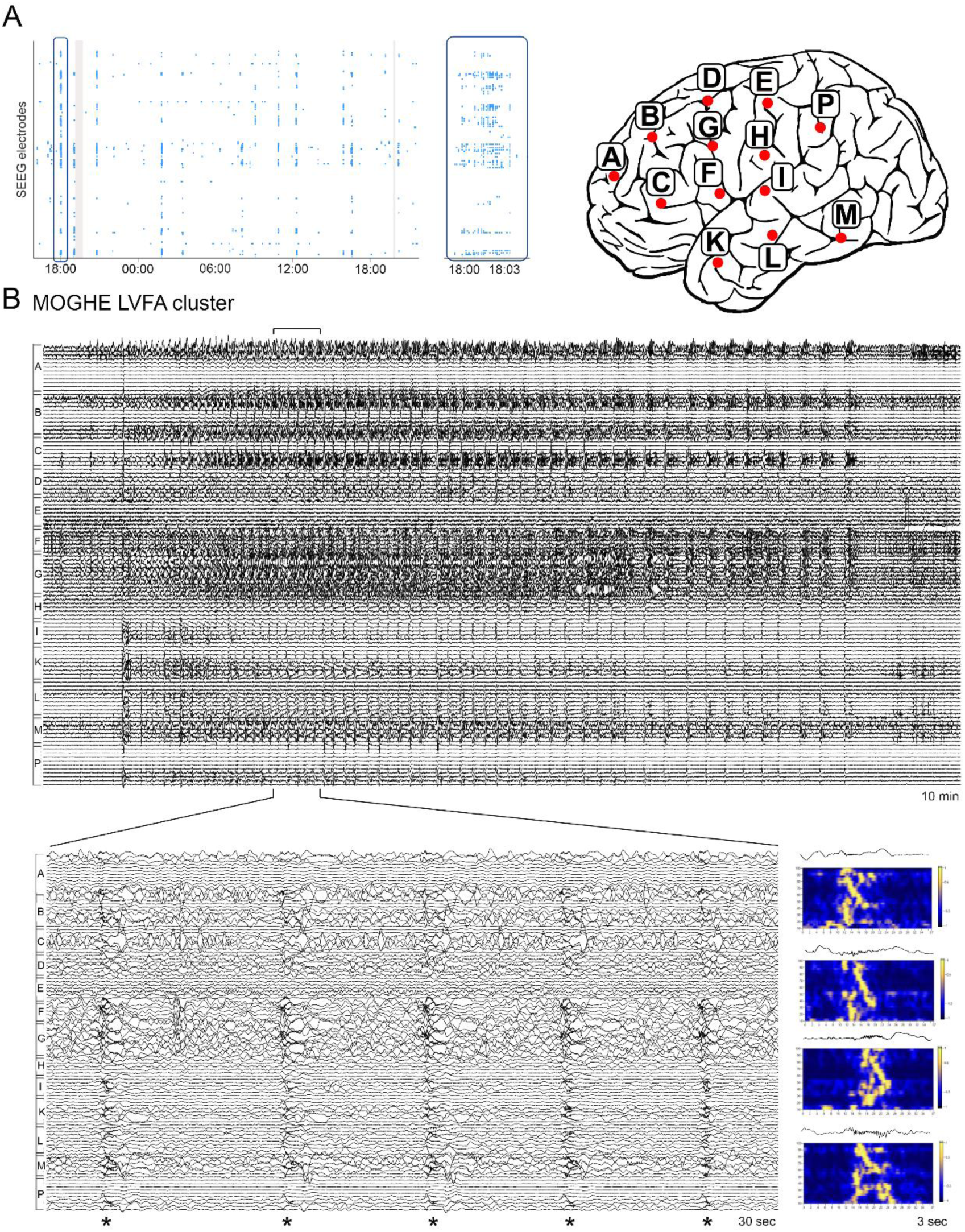
MOGHE LVFA cluster dynamics. (**A**) Overview of clustered LVFA occurrence in intracranial EEG recording over a select 24h time window shown as a raster plot (left panel). Inset highlighting a brief ∼3 min cluster. Corresponding implantation scheme of the same patient demonstrating depth electrode coverage of the frontal, temporal, and parietal lobes (right panel); the contralateral implantation is omitted for clarity. (**B**) Ten-minute intracranial EEG segment illustrating a prolonged “status-like” LVFA cluster in the same patient with evolving temporal dynamics involving widespread recruitment of the sampled epileptic network across frontal, temporal, and parietal regions. The inset shows a 30-second high-resolution segment containing five stereotypic rhythmic LVFA events from the cluster. Corresponding power spectral analysis demonstrates the characteristic chirp-like frequency pattern associated with individual LVFA events.

Repetitive LVFA clusters were observed in 16 of 18 MOGHE patients and in none of the non-MOGHE cases (Fisher’s exact test, *p*=2.43e-9), yielding a sensitivity of 88.9% (95% CI 65.3-98.6%), specificity of 100% (84.6-100%), positive predictive value of 100% (79.4-100%), and a negative predictive value of 91.7% (73.0-99.0%). Formal electroclinical correlation analyses were not performed, and synchronized video recordings were not available for systematic review. Nevertheless, clusters of epileptic spasms were frequently reported by treating clinicians in MOGHE patients. The relationship between these clinical events and the recurrent LVFA clusters identified on intracranial EEG could not be assessed in the present study.

These findings identify repetitive LVFA clusters as a distinct iEEG pattern in MOGHE with potential diagnostic value. The pattern occurred during both ictal and non-ictal periods, suggesting that the usual separation between ictal and interictal activity does not fully capture the electrophysiological behavior of MOGHE.

### Validation cohort: The iEEG pattern in MOGHE is highly specific

To assess the specificity and diagnostic utility of the identified MOGHE iEEG pattern, we performed a blinded analysis of intracranial EEGs from an independent validation cohort comprising 22 patients, equally divided between MOGHE and non-MOGHE lesions. The histopathological diagnosis was concealed from the EEG evaluator (VG), who had access only to implantation schemes and complete intracranial EEG recordings (mean recording duration, 6 days; **Table 1**). Using the same visual and computational analysis pipeline as in the test cohort, we found LVFA clusters in 10 out of 11 MOGHE cases and in 1 out of 11 non-MOGHE cases (Fisher’s exact test, *p*=0.00036), corresponding to a sensitivity and specificity of 90.9% (CI 58.7-99.8%) each. Agreement between iEEG-based prediction and histopathological diagnosis was near perfect (Cohen’s κ = 0.82).

Across the combined cohort (n=60), clustered LVFA remained most strongly associated with MOGHE histology. In multivariable ridge-penalized logistic regression with 10-fold cross-validation, simultaneously adjusting for histology, frontal versus non-frontal localization, age at seizure onset, and age at implantation, MOGHE histology remained the dominant independent predictor of clustered LVFA (adj. OR 398.87, bootstrap 95% CI 76.31-1689.93; **Fig. 4A**), whereas frontal localization showed no independent association after adjustment (adj. OR 0.09, 95% CI 0.02-2.01). Older age at seizure onset remained negatively associated with clustered LVFA (adj. OR 0.16, 95% CI 0.04-0.57), while age at implantation was not independently significant (adj. OR 0.75, 95% CI 0.15-1.62). Model discrimination remained excellent, with an apparent ROC AUC of 0.991 and a 10-fold cross-validated ROC AUC of 0.977 (**Fig. 4B**). Multivariable Firth bias-reduced logistic regression, simultaneously adjusting for histology, frontal versus non-frontal localization, age at seizure onset, and age at implantation, yielded similarly strong associations, confirming robustness (**Supplementary Table 1**).

**Figure 4:**
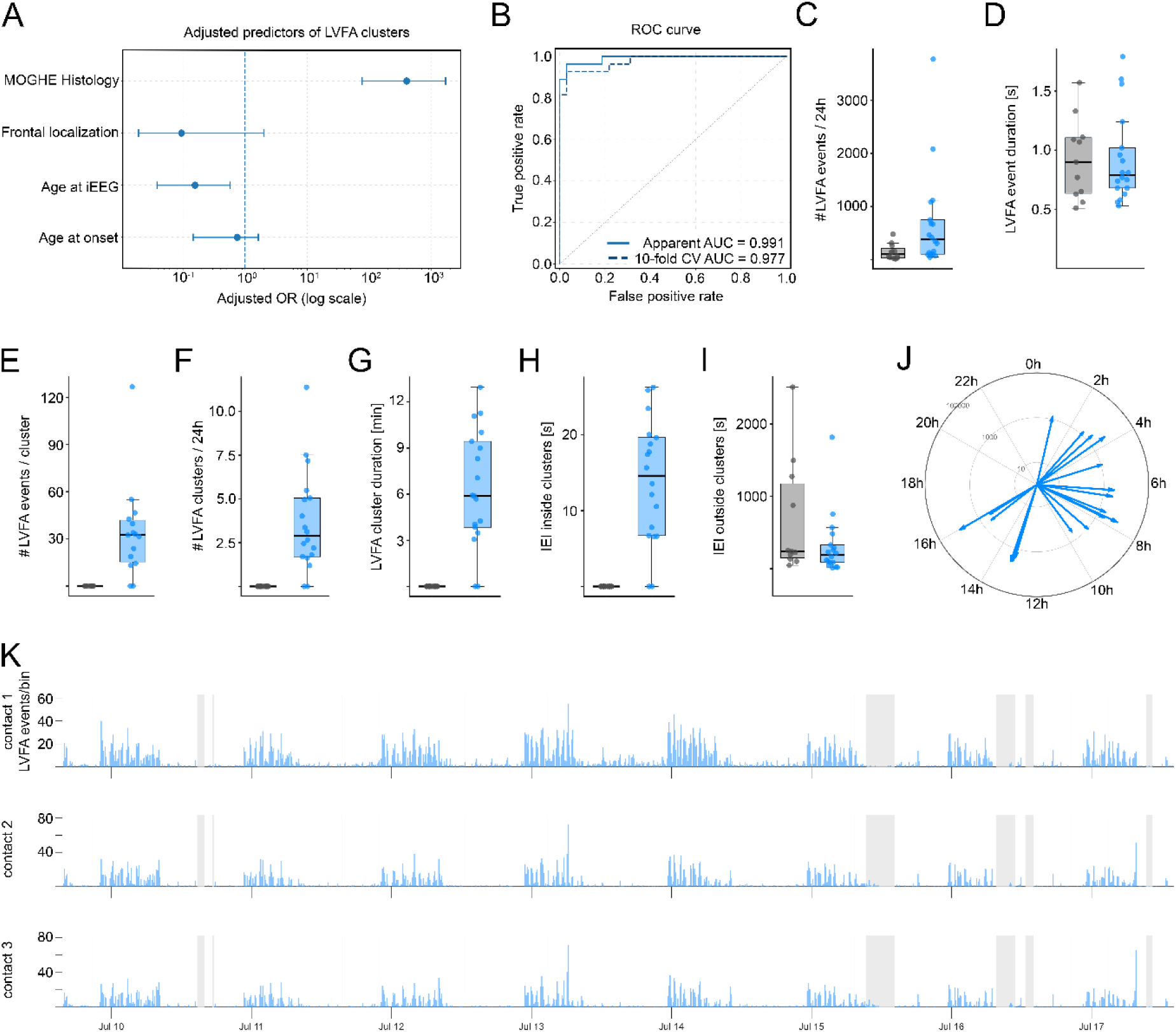
MOGHE LVFA cluster quantitative evaluation. (**A**) Forest plot showing adjusted odds ratios with bootstrap-derived 95% confidence intervals for clinical predictors of clustered LVFA obtained from ridge-penalized multivariable logistic regression. (**B**) Receiver operating characteristic (ROC) curve demonstrating discriminatory performance of the multivariable model for the prediction of clustered LVFA. (**C-I**) Quantitative characteristics of LVFA events and clusters in MOGHE (blue) and non-MOGHE patients (grey), including LVFA event burden, event duration, cluster size, cluster frequency, cluster duration, and intra- and inter-cluster event intervals. (**J**) Circular phase plots illustrating patient-specific circadian organization of LVFA clusters in MOGHE, displayed as phase-locking vectors (log-scale) and resultant angles for individual patients. (**K**) Representative circadian histograms of LVFA event occurrence across top three electrode contacts with the highest LVFA burden in a representative MOGHE patient.

To determine whether the association between clustered LVFA and MOGHE could be explained by the predominance of frontal lesions or age at implantation, additional stratified frontal-only and pediatric-only (implantation age <18 years) analyses were performed. Among frontal cases, clustered LVFA was observed in 22 of 25 MOGHE patients and in none of the 14 frontal non-MOGHE cases (Fisher’s exact test, *p* = 4.51e-8; continuity-corrected OR 186.4, 95% CI 9.0-3881.3). In the pediatric subgroup, clustered LVFA was present in all 20 MOGHE patients and in none of the 10 non-MOGHE patients (Fisher’s exact test, *p* = 3.33e-8; continuity-corrected OR 861.0, 95% CI 15.9-46535.3).

Together, these findings support clustered LVFA as a highly specific electrophysiological biomarker of MOGHE in the present study cohort that remains strongly associated with histopathology after accounting for differences in seizure localization and age.

### Quantitative iEEG analysis further supports the specificity of LVFA clusters for MOGHE

We next implemented an automated detection algorithm to quantify LVFA events occurring outside electrographic seizures. Automatically detected events were subsequently reviewed manually (VG, EP) to ensure accurate classification. iEEG full-length continuous recordings of 28 patients (18 MOGHE and 10 non-MOGHE) were analyzed, with median recording durations of 5.49 days (IQR 3.08-6.58) and 4.40 days (IQR 3.38-7.04; **Table 2**), respectively.

**Table 2:**
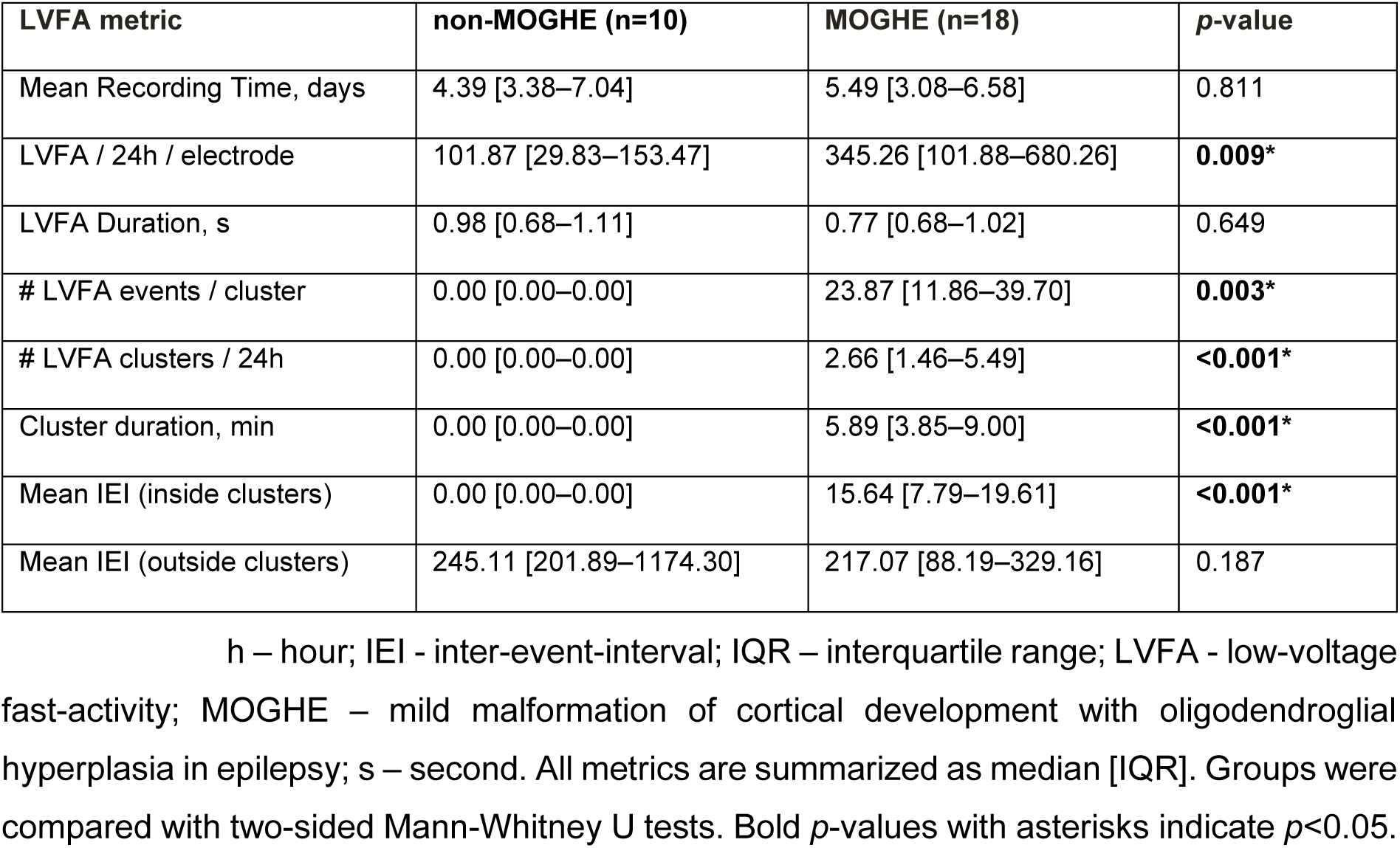
Quantitative analysis of LVFA.

Quantitative iEEG analysis further distinguished MOGHE from non-MOGHE cases with respect to both the burden and temporal organization of LVFA activity (**Table 2**). Averaged across the five electrode contacts with the highest activity, MOGHE patients exhibited substantially higher LVFA event rates than non-MOGHE patients (median 345.3 events/24h, IQR 101.9-680.3 versus 101.9 events/24h, IQR 29.8-153.5; **Fig. 4C**). In contrast, the duration of individual LVFA events was comparable between groups (MOGHE: 0.77 s, IQR 0.68-1.02; non-MOGHE: 0.98 s, IQR 0.68-1.11; **Fig. 4D**). The defining feature of MOGHE was the occurrence of prolonged repetitive LVFA clustering, which was absent in non-MOGHE cases (23.9, IQR 11.9-39.7 versus 0 events/cluster; **Fig. 4E**). MOGHE patients showed a median of 2.66 clusters/24h (IQR 1.46-5.49), with a median cluster duration of 5.89 minutes (IQR 3.85-9.00; **Fig. 4F-G**). Within clusters, LVFA events recurred at short and relatively regular intervals (median inter-event interval, 15.6 s, IQR 7.79-19.61; **Fig. 4H**), whereas inter-event intervals outside clusters were longer and more variable in both groups (245.1 s, IQR 201.9-1174.3, versus 217.1 s, IQR 88.2-329.2; **Fig. 4I**). Temporal analysis further revealed patient-specific diurnal organization of LVFA activity, with recurrent clustering phases across the 24-hour cycle in several MOGHE patients. Most MOGHE patients exhibited peak LVFA vector phases during the early morning hours, whereas a smaller subgroup showed maxima in the afternoon, potentially suggesting distinct LVFA-associated circadian chronotypes based on recurrent event and cluster timing (**Fig. 4J-K**).

Taken together, these findings indicate that MOGHE is characterized not only by the qualitative presence of LVFA clusters but also by a distinct quantitative pattern of pathological iEEG activity.

## Discussion

The present multicenter retrospective study identifies repetitive clusters of low-voltage fast activity (LVFA) as a highly specific intracranial EEG biomarker of MOGHE, extending current concepts of seizure-onset patterns in malformations of cortical development. In both the test and validation cohorts, MOGHE was characterized by stereotyped clusters of very brief LVFA events that recurred several times per minute and formed several-minute-long “status-like” episodes whose clinical correlates, however, could not be systematically assessed. These clusters occurred in 16/18 MOGHE patients in the test cohort and 10/11 MOGHE patients in the blinded validation cohort, with one false-positive association across both cohorts, yielding high sensitivity and specificity at the individual-patient level. The persistence of the association in frontal-only and pediatric-only subgroups argues against clustered LVFA being explained by age or frontal localization alone and supports its interpretation as a histology-linked electrophysiological signature of MOGHE. Despite acquisition and data heterogeneity across centers and systems, LVFA clusters were identifiable on raw bipolar traces using only standard ≥60-second review windows, arguing against a strong dependence on system-specific high-frequency sensitivity and making the result clinically actionable.

LVFA at seizure onset itself was not specific to MOGHE and occurred in all lesion types, consistent with prior work showing LVFA as the most frequent seizure-onset pattern in focal epilepsies and particularly in malformations of cortical development and focal cortical dysplasia ^19; 20; 22–26^. Thus, MOGHE is not defined by the presence of LVFA, but by its temporal organization into repetitive, network-wide clusters that recur over days and blur conventional ictal-interictal boundaries. This meso- to macro-temporal behavior is not captured when iEEG is examined only in small snippets around individual clinical seizures, underscoring the value of extended time-scale review.

An important unresolved question is whether LVFA clustering is equally prevalent in patients with the milder MOGHE phenotype characterized by later seizure onset and preserved cognition ^1^. Such patients were underrepresented in our cohorts. Notably, multivariable analysis identified younger age at seizure onset as an independent predictor of clustered LVFA, suggesting that the pattern may be enriched in early-onset forms of MOGHE. However, formal electroclinical correlations were not performed, and the present study was not designed to determine whether LVFA clustering varies across the clinical spectrum of MOGHE.

LVFA is now well established as a core seizure-onset pattern in stereo-EEG, often associated with a more localized epileptogenic zone and better postsurgical seizure control when adequately resected ^23; 27^. Mechanistic studies further suggest that LVFA seizures are associated with ictal desynchronization, altered functional connectivity, and recruitment of inhibitory interneurons ^28^, underscoring that LVFA reflects a specific network state rather than a purely “fast” oscillation. The LVFA clusters we describe in MOGHE may therefore arise from a chronically active LVFA-generating substrate in lesional white matter. Multivariable modeling further demonstrated that younger age at seizure onset was independently associated with clustered LVFA, whereas frontal localization did not retain significance after adjustment for histopathology. This finding suggests that clustered LVFA preferentially emerges in immature developmental epileptic networks but is not solely explained by lobar anatomy. From a histopathological perspective, this raises the question of which lesion components are truly specific to the LVFA-cluster pattern. Heterotopic neurons in deep white matter are also present in mMCD, and myelination abnormalities occur in FCD2B, yet neither lesion type exhibits clustered LVFA, suggesting that these features alone are insufficient. In contrast, MOGHE is characterized by extensive, sometimes multilobar oligodendroglial hyperplasia with an immature myelinating oligodendroglial compartment ^4^, which may represent the critical substrate underlying the distinctive LVFA-cluster phenotype.

In addition to their characteristic spatial distribution, LVFA clusters in MOGHE also exhibited a marked temporal organization. We observed recurrent daily patterns of LVF activity, with clustering of LVFA vector maxima into distinct early-morning and afternoon phases, suggesting that MOGHE-associated LVF activity may follow patient-specific circadian chronotypes. Similar circadian organization has previously been described for seizure occurrence in focal epilepsies and likely reflects interactions between intrinsic cortical excitability, sleep-wake regulation, and network synchronization ^29–31^. The relatively stable recurrence of LVFA clusters within individual patients further supports the concept that these events are not randomly distributed over time but are embedded within temporally structured epileptic network dynamics. Whether distinct LVFA chronotypes represent reproducible neurophysiological subtypes and correlate with clinical variables such as seizure semiology, lesion localization, sleep architecture, or treatment response will require confirmation in larger, independent cohorts.

Clinically, MOGHE frequently mimics FCD on MRI and is often misdiagnosed, particularly in older patients in whom age-dependent laminar T2/FLAIR abnormalities at the corticomedullary junction become subtle ^6; 8^. In our series and in previous cohorts, most patients were initially evaluated as suspected FCD, with only FDG-PET hypometabolism reliably pointing to an epileptogenic lesion when MRI was non-specific or negative. In this context, an intracranial biomarker that distinguishes MOGHE from FCD2 and mMCD is of immediate presurgical relevance. LVFA clusters were identifiable on conventional intracranial EEG recordings across different recording systems using standard bipolar montages and longer (≥60 s) review windows, without the need for proprietary high-frequency software. This makes the biomarker directly applicable in routine stereo-EEG and grid/strip evaluations. Time-frequency analysis primarily served as an objective and time-efficient tool for detection and quantification, rather than a prerequisite for recognizing the pattern.

Our findings further highlight the importance of adequate spatial sampling during intracranial EEG evaluation. In FCD, high densities of LVFA events may be observed within a small number of contacts at seizure onset, but none of the non-MOGHE cases in our cohort exhibited the prolonged repetitive clustering pattern characteristic of MOGHE. In contrast, LVFA clusters in MOGHE were associated with progressive recruitment of increasing numbers of contacts over time, often involving large portions of the sampled network. Broader stereo-EEG coverage is therefore more likely to capture the full spatial extent of this phenomenon and may provide a more accurate representation of the underlying epileptogenic substrate. These observations are consistent with previous clinical experience showing that, despite often focal seizure onset, favorable postsurgical outcomes in MOGHE frequently require extensive resections extending beyond the immediate seizure-onset zone. The widespread and dynamic network recruitment observed during LVFA clusters may provide a potential electrophysiological explanation for this phenomenon. From a clinical perspective, early recognition of the MOGHE-specific LVFA pattern may therefore be particularly valuable during presurgical evaluation, as it could facilitate timely identification of patients in whom a more extensive epileptogenic network should be considered during surgical planning. This may be especially relevant in pediatric patients, in whom extensive resections are most commonly performed and where delays in achieving seizure control may have lasting developmental consequences.

Our data integrate naturally with the emerging molecular and clinical framework of MOGHE. Up to 50% of patients carry brain-somatic SLC35A2 variants ^10; 11; 32^, and a high incidence of Y-chromosome mosaicism, with clear genotype-phenotype correlations, has been described ^4;33^. Together with age-dependent MRI features ^8^, stereotyped histology ^5; 18^, recurrent LVFA clustering, and early-onset drug-resistant focal epilepsy or developmental and epileptic encephalopathy ^1–3^, these findings argue for a syndromic conceptualization of MOGHE rather than a purely descriptive histopathological lesion. In the future, recognizing LVFA clusters preoperatively may prompt targeted histopathological and molecular work-up, enable precise counseling on prognosis ^34^, and identify candidates for adjunct galactose supplementation ^9^ or other mechanistically informed treatments.

Several limitations define priorities for future work. Although our cohorts are among the largest systematically studied MOGHE series to date, the overall sample size, particularly for the validation cohort, remains modest, and some clinical subgroups (older patients, extra-frontal lesions) were underrepresented. Controls were restricted to mMCD and FCD2. Future studies should evaluate LVFA clustering across a broader spectrum of epileptogenic lesions, including FCD1A and other large MCD, brain tumors, and acquired lesions, to further establish diagnostic specificity. In general, meaningful electroclinical correlations in MCD critically depend on accurate histopathological classification, which often requires specialized neuropathological expertise. Notably, all MOGHE cases in this study underwent centralized histopathological evaluation or re-evaluation at the Erlangen reference center, providing a uniform diagnostic framework for interpretation of the observed EEG patterns. Formal electroclinical correlations were beyond the scope of this study, and synchronized video recordings were not available for systematic review. Although clusters of epileptic spasms were frequently reported by treating clinicians, their relationship to recurrent LVFA clusters remains unknown. Consequently, it is unclear whether LVFA clusters represent the electrophysiological substrate of these clinical events, evolving “micro-status” episodes, or a distinct pathological network state characterized by repetitive LVFA generation. Prospective studies combining long-term stereo-EEG, scalp EEG, quantitative network analysis, and standardized semiology will be required to address these questions. Inter-rater reproducibility of LVFA cluster identification and robustness to preprocessing parameters (band, threshold, montage) were not assessed and warrant future investigation. Finally, we did not assess whether the presence, burden, or diurnal organization of LVFA clusters (chronotype) predicts postsurgical outcome beyond traditional markers such as histopathology/lesion type and completeness of resection. Future multicenter studies with harmonized recording and analysis protocols should determine whether LVFA clusters improve outcome prediction and whether they remain stable or change in response to surgery and targeted therapies. If validated, LVFA clustering could become a simple, visually accessible biomarker that bridges intracranial electrophysiology with genetics, imaging, and precision treatment strategies in MOGHE.

## Acknowledgment

KK and IB are supported by the DFG, project 460333672 CRC1540 EBM.

## Ethical Publication Statement

We confirm that we have read the Journal’s position on issues involved in ethical publication and affirm that this report is consistent with those guidelines.

## Data Availability Statement

The intracranial EEG and clinical datasets analyzed in the current study contain sensitive patient information, are subject to institutional and national data protection regulations, and cannot be made publicly available.

## Author contributions

KK planned, organized, and supervised the study, and drafted the main manuscript and figures. VG and EP performed all qualitative and quantitative intracranial EEG analyses and statistics, generated graphs and figure panels, and assisted in drafting the manuscript. MK, TK, DD, VB, and KR performed surgical implantations of grids and depth electrodes. TH, AG, TC, FGW, SG, HH, SR, FM, RT, AGN, KAK, and RS handled patients included in the present study, provided clinical data, and/or helped interpret results. RC, IB, VZ, AM, and VAQ performed histopathological and genetic evaluations of patients. All authors edited and approved the final version of the manuscript.

## Declaration of generative AI and AI-assisted technologies in the manuscript preparation process

During the preparation of this work, the author(s) used ChatGPT and q.e.d.science in order to edit the manuscript for grammar, spelling, and clarity and flow, as well as for identifying conceptual or logical gaps in our narrative. The tools were not used to generate scientific content. After using these tools/services, the authors reviewed and edited the content as needed and take full responsibility for the published article’s content.

## Supplements

**Supplementary Table 1:**
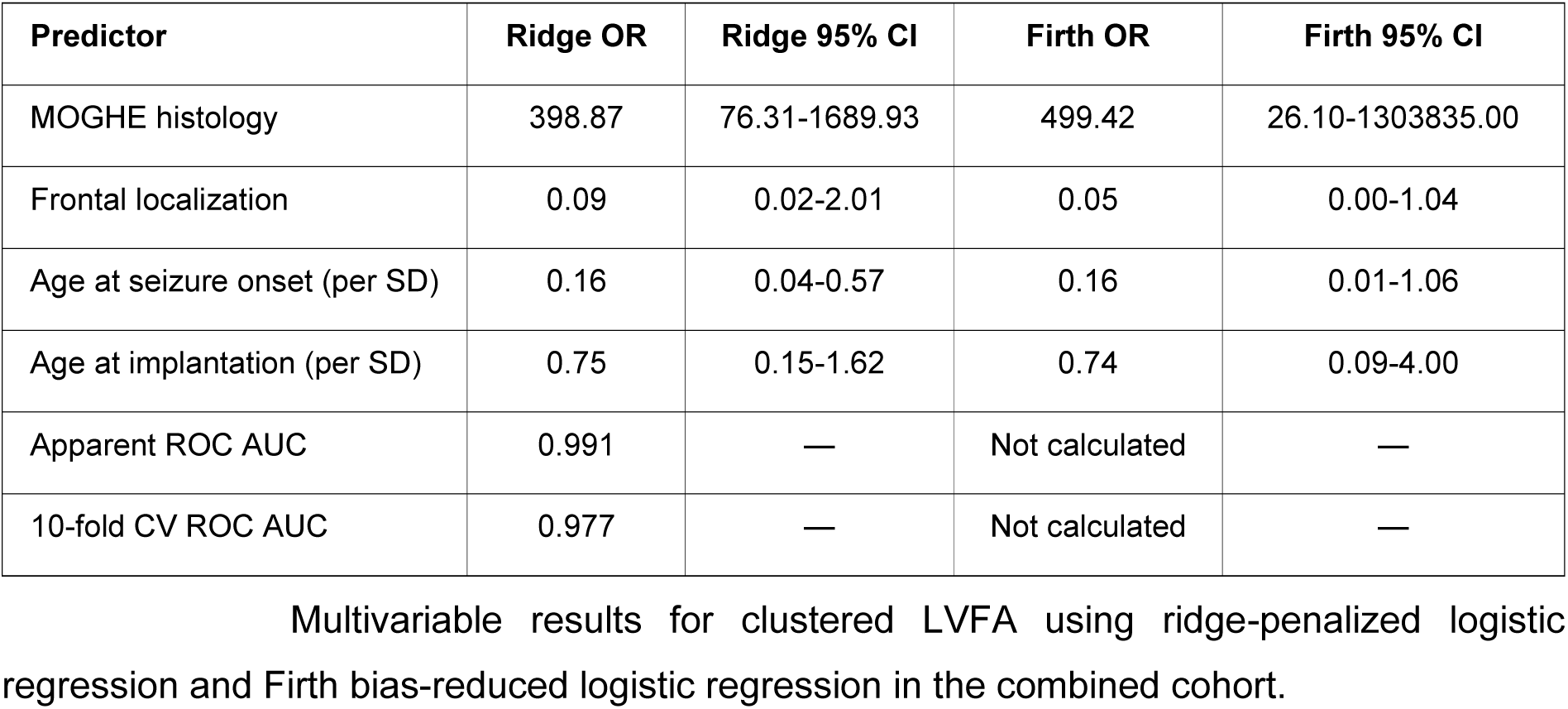
Model comparison.

